# The association between heterosexual anal intercourse and HIV acquisition in three prospective cohorts of women

**DOI:** 10.1101/2022.09.07.22279674

**Authors:** Romain Silhol, Ashley Nordsletten, Mathieu Maheu-Giroux, Jocelyn Elmes, Roisin Staunton, Branwen Owen, Barbara Shacklett, Ian McGowan, Kailazarid Gomez Feliciano, Ariane van der Straten, Leigh Anne Eller, Merlin Robb, Jeanne Marrazzo, Dobromir Dimitrov, Marie-Claude Boily

## Abstract

Receptive anal intercourse (RAI) may substantially increase HIV acquisition risk per sex act compared to receptive vaginal intercourse (RVI). To understand how levels of RAI change over time and evaluate the impact of exposure definitions for RAI on HIV incidence, we analysed three prospective HIV cohorts of women: RV217, MTN-003 (VOICE), and HVTN 907. At baseline 16.0% (RV 217), 17.5% (VOICE) of women reported RAI in the past 3 months and 27.3% (HVTN 907) in the past 6 months, with RAI declining during follow-up by around 3-fold. Hazard ratios, adjusted for potential confounders (aHR), indicate that reporting RAI at baseline increased HIV incidence in the three cohorts: 1.1 (95% Confidence interval: 0.8-1.5) for VOICE, aHR of 3.3 (1.6-6.8) for RV 217, and 1.9 (0.6-6) for HVTN 907. Using time-varying exposure definition slightly increased the estimated association for VOICE (aHR=1.2; 0.9-1.6), however reporting >30% RAI sex acts during VOICE follow-up was not associated with higher HIV incidence (aHR=0.7 (0.4-1.1)). Women who always reported RAI during follow-up where also at increased HIV acquisition risk. Overall, we found that precisely estimating RAI and HIV association after multiple RVI/RAI exposures is sensitive to RAI exposure definitions and may be influenced by measurement errors.

## Introduction

Systematic reviews of cross-sectional studies have shown that heterosexual anal intercourse (RAI) is prevalent worldwide, with little apparent variation by key demographic characteristics such as age (1-9). Our understanding of the contribution of RAI to HIV transmission dynamics is, however, limited by the scarcity of longitudinal data that tracks levels and persistence of RAI over time and the quality of RAI measurement. The probability of HIV transmission through one act of RAI may be up to 18 times higher than through one act of receptive vaginal intercourse (RVI) (10), although empirical estimates are uncertain (7, 11, 12). This suggest that even infrequent RAI in a population (5-10% of all acts) could account for a substantial number of HIV infections (4, 13), which could influence the impact of prevention strategies such as vaginal microbicide or oral PrEP, with varying efficacy by anatomical site (14-16). Nevertheless, HIV trials and cohort studies alike have typically given minimal considerations to the impact of RAI practice on HIV incidence during follow-up. A recent systematic review collating all longitudinal HIV studies that included some measure of RAI estimated that women reporting RAI were more likely to acquire HIV than women not reporting RAI (17). However, the review highlighted gaps due to many different measures of RAI exposures being used across studies (including those only measured at baseline), the near systematic use of non-confidential interview methods, lack of adjustment for important confounders such as condom use, leading to a pooled adjusted increase in risk higher than a crude estimate (2.2-fold vs 1.6-fold, respectively), and scarce information quantifying the extent to which RAI increase HIV risk if practiced frequently or unfrequently. Moreover, just a quarter of estimated associations accounted for changes in RAI practices over time which should influence the estimated magnitude of HIV risk (17).

Existing knowledge on the prevalence of RAI activity in a population have largely been based on reports of prevalence over long recall periods (lifetime, past year) (8), with frequent inconsistencies between estimates also influenced by the use of non-confidential interview methods, such as face to face interviews and/or challenges with accurate translation of sexual terms (18). However, very few cohorts looked at the prevalence and persistence of RAI over time (10, 19, 20). For example, a systematic review of the practice of heterosexual anal intercourse in South Africa has found that only one among 31 studies reporting RAI prevalence has reported it over two different time frame (3). In the review looking at the increased HIV incidence due to RAI exposure, only a quarter of the 18 reported estimates accounted for changes in RAI practices over time and less than a quarter were adjusted for possible confounders (17).

To address this knowledge gap, we used longitudinal data from three recent cohort studies conducted in South and Eastern Africa (MTN-003 (VOICE) trial and RV 217) and in the Caribbean (HVTN 907) (21-23) to 1) examine the level and persistance of RAI practice (e.g., prevalence) among women over the study periods and to 2) assess the relationship RAI and HIV incidence using different RAI exposure definitions.

## Methods

### Description of the three longitudinal studies (VOICE, RV 217, and HVTN 907)

We analysed data from three recent longitudinal studies: the MTN-003 Vaginal and Oral Interventions to Control the Epidemic (VOICE) trial (24), the prospective study of acute HIV infections in adults (RV 217)(25), and the prospective cohort study of Caribbean female commercial sex workers at high risk of HIV infection (HVTN 907)(26). The VOICE and RV 217 were conducted in south and east Africa (VOICE: South Africa, Uganda, and Zimbabwe; RV 217: Uganda, and Kenya), whilst HVTN 907 was conducted in the Caribbean (Haiti, Dominican Republic, and Puerto Rico). The main characteristics of these studies are summarised in Table 1, while their complete description, including protocols, can be found elsewhere (24-26). Briefly, VOICE recruited 5,029 sexually active women aged 18-40 years who reported at least one act of RVI in the three months prior to the baseline interview. For VOICE, we used the data from every trial arm because none of them were associated with lower HIV incidence due to low adherence. RV 217 recruited women with higher risk profiles aged 18-47 years (N=1,545) from locations associated with transactional sex (e.g. bars and clubs). HVTN 907 recruited female sex workers (FSWs) aged 18-45 years (N=1,019) who had performed at least one act of RVI or RAI without condoms in the last six months. During follow-up, behavioural data was collected every three months via *Audio Computer-Assisted Self-Interview* (ACASI) for VOICE, every six months via ACASI for RV 217, and every six months via face-to-face interview for HVTN 907. RAI practice at baseline and follow-up surveys covered RAI in the past 3 months for VOICE and RV 217, and in the past 6 months for HVTN 907 (Figure S1). Information on the number of RAI acts during these recall periods was available for VOICE and HVTN 907 participants (Tables 1 and S1). Condom use at last RAI was available for VOICE, at last RAI for 3 types of partners (steady, casual, client) for RV 217, but not collected during HVTN 907. Although women enrolled in RV 217 were followed every six months, the behavioral questionnaire used a recall period of three months (Figure S1), and we assumed that the participants’ report of RAI practice over their past six months was the same as over the past 3 months.

**Table 1.** Overview of HVTN 907, VOICE, and RV 217 studies’ characteristics, including a description of key demographic and behavioural participant characteristics.

|  | **VOICE**<br>**(Southern Africa)** | **RV 217**<br>**(East Africa)** | **HVTN-907**<br>**(Caribbean)** |
| --- | --- | --- | --- |
| **Study overview:** |  |  |  |
| Years of data collection | 2009-2012 | 2009-2018 | 2009-2012 |
| Site | South Africa, Uganda,<br>and Zimbabwe | Uganda<br>and Kenya | Haiti, Dominican Republic,<br>and Puerto Rico |
| Study type | Randomized trial of oral and<br>gel pre-exposure prophylaxis<br>(PrEP) | Observational study | Observational study |
| Sample Size (baseline) | N=5,029 | N=1,545 | N=1,019 |
| Data Method | Audio computer-assisted<br>self-interview (ACASI) | Audio computer-assisted self-interview<br>(ACASI) | Face-to-face interview<br>(FTFI) |
| Population Type | Females at risk of HIV<br>infection | FSW (90%) &<br>high-risk females | FSW |
| Recruitment | STI/family<br>planning/postnatal clinics | Community/<br>street outreach | Street outreach |
| Age Range | 18-40 | 18-47 | 18-45 |
| Minimum Inclusion | HIV-uninfected and sexually<br>active (RVI in last 3 months) | Exchanged sex and<br>condomless RVI or<br>condomless RAI with ≥<br>3 partners or ≥ 1 HIV-positive partners | Exchanged sex plus<br>condomless RVI or<br>condomless RAI in prior 6<br>months |
| Follow-up visits<br>(behavioural) | Every 3 months for 3 years | Every 6 months for 2<br>years | Every 6 months for 18<br>months |
| Person-years of follow-up | 5,509 | 1,589 | 1,119 |
| Injection Drug Use | Excluded | Included | Included |
| HIV test window and<br>frequency of HIV testing | 1 month (rapid test)<br>Monthly | 10 days (RNA tests)<br>Twice weekly | 1 month (normal test)<br>Every 6 months |
| HIV Incidence<br>(seroconversions during<br>follow-up) | 306 infections | 34 infections | 12 infections |
| **Receptive anal intercourse (RAI) and demographic variables:** |  |  |  |
| RAI Variables | RAI in last 3 months<br>Number (No.) of RAI acts in<br>last 3 months | RAI in last 3 months<br>RAI in last 3 months<br>per partner type<br>No. RAI<br>partners/partner type<sup>a</sup> | RAI in last 6 months<br>No. of RAI acts in last week |
| Condom use during RAI | During last RAI | During last RAI with<br>each partner type<sup>a</sup> | Only the number of<br>condomless RAI in prior<br>week was reported |
| Age categories | <25, 25+ years | <25, 25+ years | <25, 25+ years |
| Trial arms | All<sup>b</sup> | NA | NA |
| Ethnicity | NA | NA | Hispanic/Black/White/Other |
| Injection Drug Use | NA | IDU history | IDU and non-IDU history |
| **Other sexual behaviours:** |  |  |  |
| Sex work | Prior year | Prior 3 months | Prior 6 months |
| Vaginal intercourse<br>variables | No. RVI in prior week | No. acts (not reported<br>if RAI or RVI) in prior 3<br>months, by partner<br>type<sup>a</sup> | Only the number of<br>condomless RVI in prior<br>week was reported |
| Condom use during RVI | Last RVI in last week | Last intercourse (not reported if RAI or RVI),<br>by partner type<sup>a</sup> | NA (see above) |

RAI= receptive anal intercourse; IDU=injecting drug users; NA=not available; PrEP=pre-exposure prophylaxis; STI=sexually transmitted infection; RVI=vaginal intercourse;
<sup>a</sup> Four partner types were defined during the RV217 study: 1) steady partners, 2) casual partners, 3) client (commercial sex partners), and 4) “other” partners (which includes boss/work supervisor, teacher, other authority figure and relative other than spouse, student, employee/work subordinate, or rapist)
<sup>b</sup> Every trial arms were included for this analysis because none of them were associated with higher HIV incidence(24).

Incident HIV infections were measured by testing participants once a month in the VOICE trial using two different third generation rapid tests (positive assays were confirmed by a GS HIV-1 western blot)(24) and every six months in the HVTN 907 study using ELISA tests (26). HIV-negative women enrolled in RV 217 were tested twice a week using Aptima RNA tests on small-volume blood samples(25).

### Persistence of RAI practice over time

In our analysis, RAI prevalence was defined as the proportion of the study population that reported having practiced RAI over a specific recall period and RAI fraction as the fraction of total sex acts (RVI plus RAI) that were anal, only among those that reported this sexual behaviour.

To characterize the RAI practices among study participants, we derived five outcomes which describe RAI patterns over follow-up time: 1) cross-sectional RAI prevalence at baseline and 2) at each subsequent follow-up visit, 3) anytime RAI prevalence, 4) the proportion of women reporting RAI for the first time and 5) the proportion of women who stopped practicing RAI during follow-up.

Cross-sectional RAI prevalence was defined as the proportion of women reporting RAI over the previous 3 months (6 months for HVTN) at the relevant baseline or follow-up visit, whereas anytime RAI prevalence was defined as the proportion of women that reported RAI at baseline or any of the follow-up visits. Both outcomes were also stratified by participant socio-demographic characteristics. Total changes in RAI prevalence during the study was evaluated by comparing RAI prevalence at first (baseline) and last visit. These changes were investigated using chi-squared test for linear trend in proportions.

The proportion of women ceasing RAI practice was estimated from the prevalence at first follow-up visit, but only among reporting RAI at baseline (hereby called “*RAI+ women*”). Women initiating RAI was estimated from the prevalence at first follow-up, but only among women reporting only RVI at baseline (hereby called “*RVI-only women*”). Anytime RAI was defined as the proportion of participants that reported RAI at some point of baseline or follow-up. Finally, we derived the RAI fraction of sex acts for the VOICE and HVTN 907 studies at each visit (RV 217 did not collected RAI frequency data) that were anal among women reporting RAI, as well as the cross-sectional fraction of condom use during RVI and RAI for VOICE and RV 217. The prevalence of condomless RAI could not be ascertained as information on condom use during RAI was only recorded at last RAI for VOICE and RV 217, and not collected during HVTN 907 (see studies descriptions section).

### RAI HIV incidence ratio at baseline and during follow-up

Our three studies tested participants for HIV at varying frequencies and the tests had diverse detection window periods. To address these issues, dates of HIV infection were inferred in different ways. The seroconversion time was assumed at mid-point between the last and current visit for VOICE and HVTN 907, and HIV infections of participants were assumed to have occurred one month before seroconversion time (Figure 1 for VOICE). For participants of RV 217, we assumed that HIV infection occurred two weeks before the first positive HIV test, reflecting the high testing frequency and very short detection window of HIV RNA tests. Only 12 seroconversions were observed in the HVTN 907 study, thus the association between RAI practice and HIV was calculated as the ratio of cumulative incidences among RAI+ and RVI-only women, and only the VOICE trial and the RV 217 study were used for detailed analyses.

**Figure 1.**
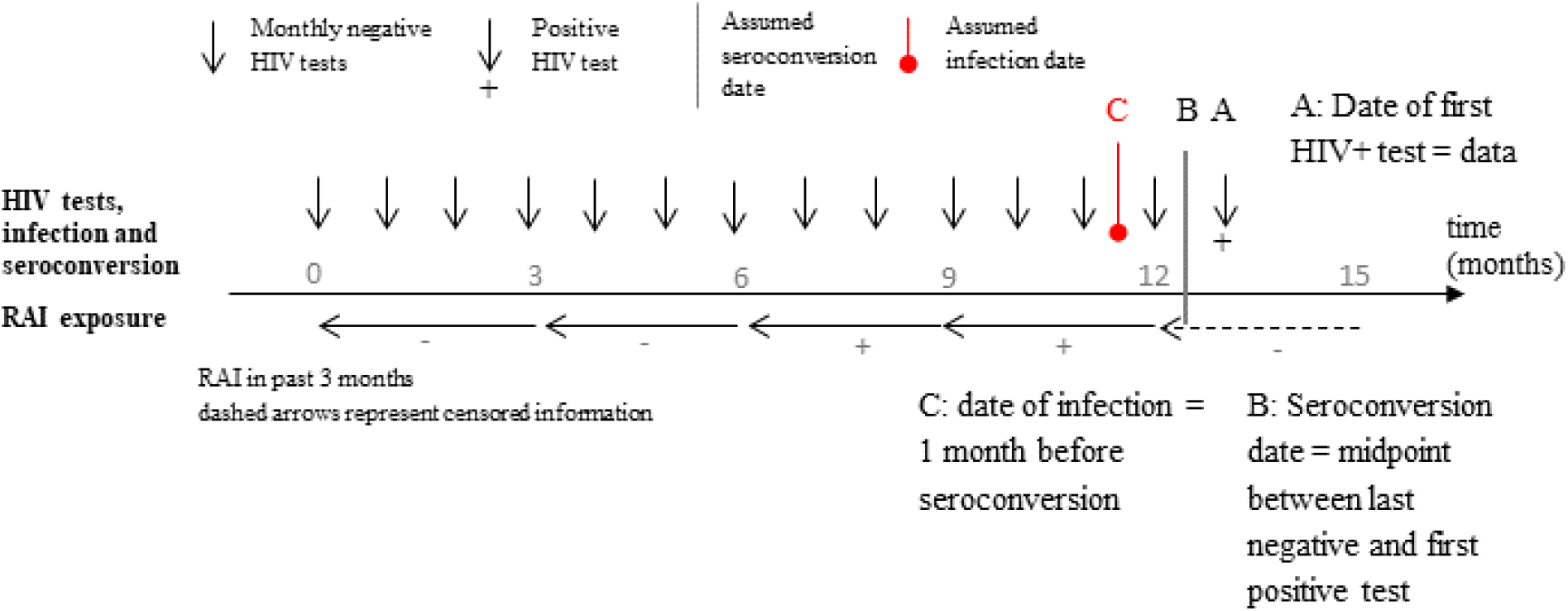
Inferring the date of HIV infection (C) among VOICE participants. In this example, the behavioural data collected after 15 months (dashed arrow) was not used because the covered period was after the estimated infection time. The plausible exposure to receptive anal intercourse (RAI) considered in the analysis is represented by “+”.

Five different RAI exposure definitions (D1-D5) commonly used in previous studies were examined in this study. The first one consisted as having reported RAI at baseline (D1; in the past three months for VOICE and RV 217). The second definition reflected RAI persistence during follow-up visits; each woman was classified as having never or ever reported RAI during follow-up (D2). The third RAI exposure definition classified participants as never, sometimes (D3_a_), or always (D3_b_) reporting RAI during follow-up (excluding baseline information). The fourth RAI exposure definition was time-varying based on reports of RAI practice at each visit (D4; baseline and follow-up) (27, 28). The last definition was based on the RAI fraction (for VOICE trial only) and was expressed as the proportion of all sex-acts that were RAI among those reporting RAI during follow-up (D5_a,b_).

The increase in HIV risk associated with RAI practice was first estimated using univariate Cox Proportional hazards models. Analyses were performed separately for the VOICE and RV 217 studies due to differences in populations and study designs. Potential confounders of the RAI-HIV relationships were then adjusted for in multivariable analyses using available data from each study. For the VOICE trial, the multivariable model was adjusted for baseline data on age at enrolment (18-25, 25+ years), country (South Africa, Uganda, or Zimbabwe), trial arm (control vs. placebo), sex work (prior year), number of partners (1, 2, 3+ in the past 3 months), and condom use at last vaginal sex. The adjusted model for the RV 217 study included: age (18-25, 25+ years), country (Uganda or Kenya), number of partners in the last 3 months (<10, 10+), history of injecting drug use (never/ever), and condom use at last sex. None of the model could be adjusted for condom use at last RAI because it was similar with condom use at last RVI in VOICE (69% vs 71%) or last sex by partner type in RV 217 (e.g. 47% vs 53% at last sex with a client), and only reported by RAI+ women.

Two different versions of the ACASI questionnaire were used during the VOICE trial, reflecting improved translation of terms related to RAI practice. A sensitivity analysis evaluated the impact of gradually introducing this new questionnaire on the persistence of RAI by comparing the cross-sectional RAI prevalence for each questionnaire version, and on the estimates of the association between RAI exposure definitions and HIV incidence. All statistical analyses were performed using the R software (version 3.5.1) (29).

## Results

### Prevalence, frequency, and persistence of RAI practice in the three longitudinal studies

The proportion of women reporting RAI over the last three months at baseline were 17.5% (95% confidence interval (95%CI): 16.5-18.6%) among VOICE participants and 16.0% (14.2-17.9%) among RV 217 participants (Figure 2). For HVTN 907, prevalence of RAI over the past six months was 27.3% (24.4-30.2%).

**Figure 2.**
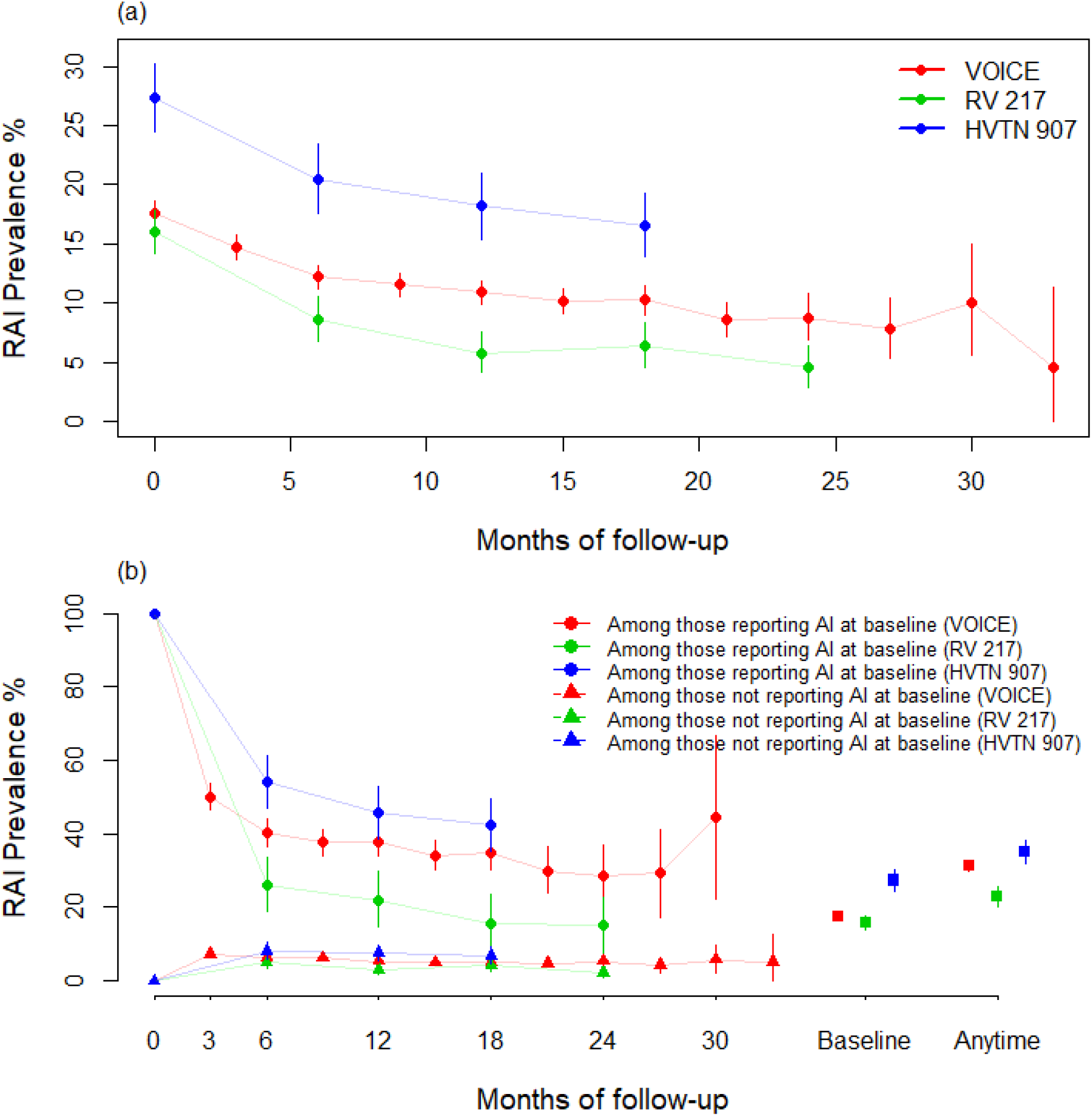
(a) Cross-sectional prevalence of receptive anal intercourse (RAI) at baseline and during follow-up for the three longitudinal studies under consideration (VOICE, RV 217, and HVTN 907). (b) Cross-sectional RAI prevalence among women that reported RAI at baseline (dots). The same, but calculated only among women that did not report RAI at baseline (triangles). On the right, baseline RAI prevalence (in the past 3 months for VOICE and RV 217, 6 months for HVTN 907), and anytime RAI prevalence (correspnding to 3 years, 2 years, and 18 months of follow-up for VOICE, RV 217, and HVTN 907, respectively). Error bars represent 95%CI of data.

Cross-sectional RAI prevalence at follow-up visits significantly decreased over time in all three studies (Chi-square test for trend: p-value<0.001 for VOICE, RV 217, and HVTN 907) (Figure 2a). RAI prevalence (past 3 months) decreased by 74.3% after 33 months of follow-up in VOICE (from 17.5% at baseline to 4.5%) and by 71.9% after 2 years of follow-up (from 16.0% to 4.5%) in RV 217. RAI prevalence (over past 6 months) decreased by 39% after 18 months of follow-up in HVTN 907 (from 27.3% to 16.6%).

Results of the persistence of RAI practice stratified by socio-demographic characteristics are presented as supplementary material (Figures S2 to S6). Briefly, the decrease in cross-sectional RAI prevalence over two years was largest for Kenyan participants of RV 217 declining from 14.1% (11.8-16.2%) at baseline, to 1.5% (0.5-2.8%) after two years of follow-up (Figure S2).

Most participants did not practice RAI at any time during follow-up (74.8% VOICE, 86.8% RV 217, and 72.8% for HVTN 907) (Figure S7), while 21.9%, 11.1%, and 16.3% of participants in VOICE, RV 217, and HVTN 907 respectively reported RAI at least once during follow-up. Only few participants practiced RAI at all follow-up visits (3.3% VOICE, 2.1% RV 217, and 10.9% for HVTN 907). Among those ever-practicing RAI (25.2% VOICE, 13.2% RV 217, and 27.2% for HVTN 907), reporting RAI at only one follow-up visit was most common.

The anytime RAI prevalence during baseline and follow-up was 31.1% (29.9-32.5%) for VOICE, 23.1% (20.3-25.9%) for RV 217, and 35.3% (32.1-38.4%) for HVTN 907 (Figure 2b). Large number of women reporting RAI at baseline reporting no RAI during subsequent follow-up visit and the prevalence of RAI among these women decreased during follow-up. The decline in reported RAI practice was most pronounced between baseline and first follow-up, with 49.9% of individuals in the VOICE sample who were RAI+ at baseline no longer reporting RAI at the three-month follow-up (Figure 2b). The decrease was even steeper in RV 217; 73.7% of them who had stopped after 6 months. In HVTN 907, the reductions were 45.9% between baseline and first follow-up – also occurring at six months. The RAI prevalence among these women continued to decrease after this first follow-up visit.

Importantly, 7.2% (6.4-8.1), 5.0% (3.4-10.6), and 8.1% (5.9-10.6) of the women who did not report RAI at baseline in VOICE, RV 217 and HVTN 907 reported initiating it at the first follow-up visit (3, 6, 6 months after baseline, respectively) (Figure 2b). The characteristics of RAI+ women most associated with ceasing reporting RAI at first follow-up were residing in Zimbabwe (VOICE), Kenya (RV 217), and Haiti (HVTN 907) (Figures S4a-6a). Conversely, being aged under 25 years (VOICE), residing in Uganda (RV 217) and Puerto Rico (HVTN) were most associated with reporting initiating RAI at first follow-up visit among RVI-only women (Figures S4b-6b).

The fraction of RAI among those who reported RAI acts was 34.2% (31.6-37.1) in VOICE and 16.0% (13.7-18.5) in HVTN 907 at baseline. These proportions as well as the fraction of condom use during RVI and RAI remained stable after initial counselling and during follow-up in both studies (Figures 3 and S8, S9).

**Figure 3.**
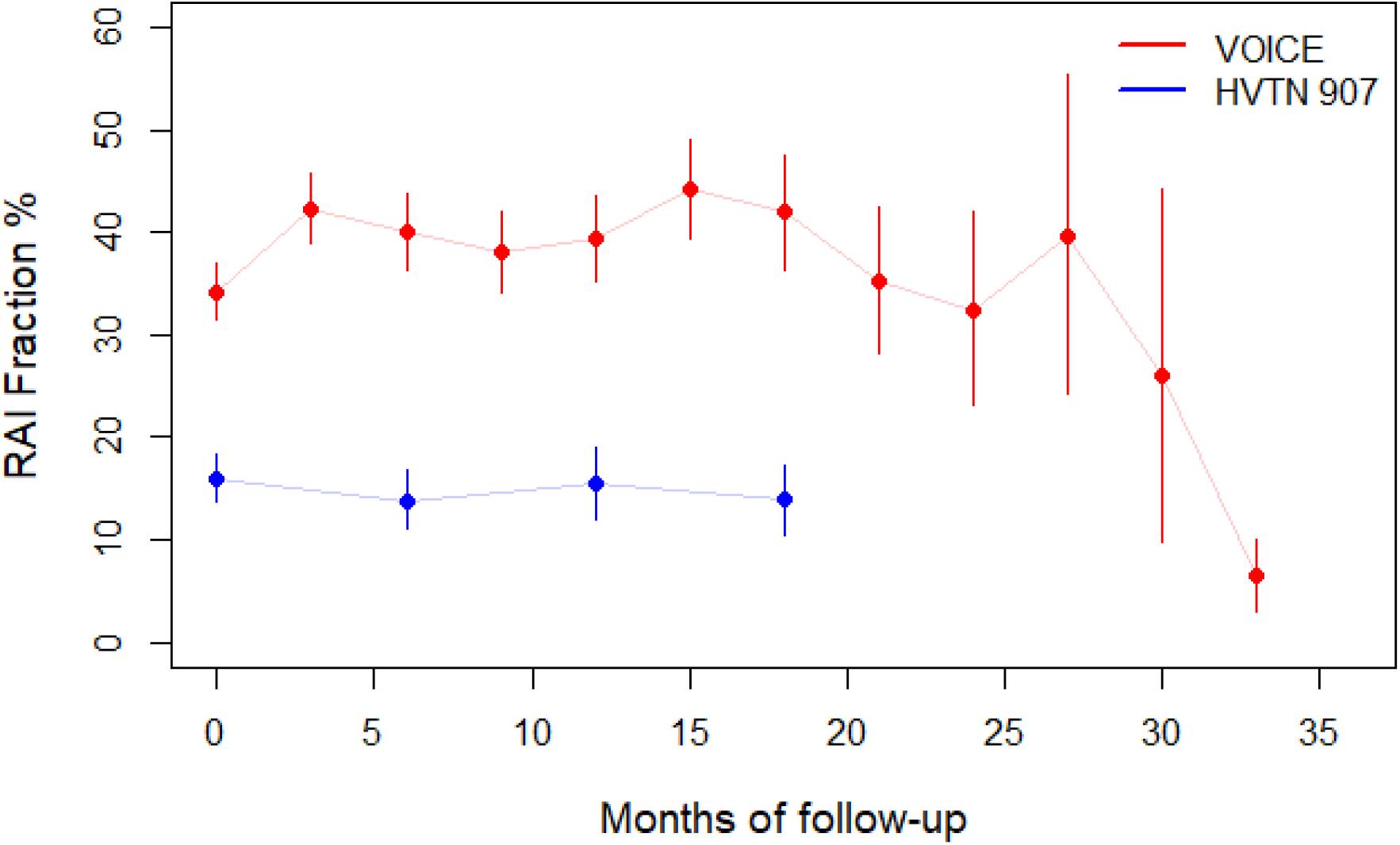
RAI fraction during VOICE (red) and HVTN 907 (blue): cross-sectional fraction of sex acts that are receptive anal intercourse (RAI), calculated among participants with positive recent history of RAI. Error bars represent 95%CI of data.

### Incident HIV and RAI association for different RAI exposure definitions

Overall, we found positive associations between the different RAI exposure definitions and incident HIV across studies, albeit accompanied by sometimes wide confidence intervals (Table 2). The RAI HIV cumulative incidence ratio for HVTN 907 was 1.9 (0.6-6), based on 12 incident cases (5 among RAI+, 7 among RVI-only women). For VOICE and RV 217, the different definitions of RAI exposure provided different levels of association with HIV infection. Reporting RAI in the three months before baseline was associated with higher HIV incidence during RV 217 (aHR=3.3; 1.6-6.8), but not during VOICE (aHR=1.1; 0.8-1.5). Reporting RAI at any time during follow-up (D2) was not associated with higher HIV incidence, although the point-estimate of association was relatively high for RV 217 (HR=1.7; 0.4-7.2). Consistently reporting RAI during follow-up (D3_b_) was associated with higher HIV incidence in both VOICE (aHR=2.0; 1.3-3.1) and RV 217 (aHR=2.6; 0.3-19.2). The unadjusted time-varying definition (D4) yielded a slightly higher point-estimate of association than D1 (1.2 vs 1.1) for VOICE. Surprisingly, the reported fraction of acts that were RAI (D5_a,b_) was not associated with HIV incidence during VOICE. We were not able to calculate adjusted hazard ratio (aHR) for D2-D4 nor use time-varying RAI variables for RV 217 due to low incidence (22 infections) among individuals completing at least one round of follow-up questionnaire.

**Table 2.** Epidemiological data used of the longitudinal analysis, and results of statistical associations between receptive anal intercourse (RAI) exposures and HIV infection during the studies, hazard ratios (HR) and adjusted hazard ratios (aHR) point estimates and 95% confidence intervals.

|  | VOICE<br>(Southern Africa) |  | RV 217<br>(Africa) |  |
| --- | --- | --- | --- | --- |
| Number of HIV seroconversions |  |  |  |  |
| Among all participants | 312 |  | 34 |  |
| Among participants completing at least one round of follow-up | 306 |  | 22 |  |
| Definitions of receptive anal intercourse (RAI) exposure | HR (95%CI) | aHR (95%CI)<sup>a</sup> | HR (95%CI) | aHR (95%CI)<sup>b</sup> |
| D1: RAI at baseline |  |  |  |  |
| No RAI | Referent | Referent | Referent | Referent |
| RAI | 1.23 (0.93-1.64) | 1.10 (0.82-1.48) | 3.26 (1.63-6.5) | 3.33 (1.64-6.79) |
| D2: RAI during follow-up (excludes baseline information) |  |  |  |  |
| Never RAI during follow-up | Referent | Referent | Referent | Referent |
| Ever RAI during follow-up | 0.91 (0.69-1.21) | 0.80 (0.60-1.07) | 1.68 (0.39-7.18) | NA |
| D3<sub>a,b</sub>: RAI frequency during follow-up (excludes baseline information) |  |  |  |  |
| Never RAI during follow-up | Referent | Referent | Referent | Referent |
| Sometimes RAI during follow-up | 0.66 (0.47-0.93) | 0.59 (0.41-0.83) | 0.34 (0.04-2.51) | NA |
| Always RAI during follow-up | 2.29 (1.51-3.47) | 2.00 (1.29-3.12) | 2.57 (0.34-19.17) | NA |
| D4: Time-varying RAI exposure |  |  |  |  |
| No RAI in preceding period | Referent | Referent | Referent | Referent |
| RAI reported | 1.33 (1.02-1.75) | 1.18 (0.88-1.57) | NA | NA |
| D5<sub>a,b</sub>: RAI fraction (fraction of acts that are RAI) |  |  |  |  |
| No RAI | Referent | Referent | Referent | Referent |
| 1-30% of sexual acts are RAI | 0.98 (0.71-1.35) | 0.86 (0.61-1.20) | NA | NA |
| >30% of sexual acts are RAI | 0.81 (0.50-1.29) | 0.69 (0.42-1.14) | NA | NA |

RAI=anal intercourse; HR= Hazard ratio; aHR= adjusted Hazard ratio; 95%CI=95% confidence interval.
<sup>a</sup> VOICE model adjusted for baseline data on age (18-25, 25+ years), country (South Africa, Uganda, or Zimbabwe), trial arm (control vs. placebo), sex work (prior year), number of partners (1, 2, 3+ in the past 3 months), and condom use at last vaginal sex
<sup>b</sup> RV 217 model adjusted for age (18-25, 25+ years), country (Uganda or Kenya), number of partners in the last 3 months (<10, 10+), history of injecting drug use (never/ever), and condom use at last sex. The later was defined as condom use at last sex with client, or with a casual partner if condom use with last client was not reported.

## Discussion

Our study shows that RAI was commonly practiced by women recruited in HIV longitudinal studies, but that this practice declined markedly during the study periods. Despite its well-established heightened per-act risk of HIV acquisition, the association between RAI practice and incident HIV infection was not always positive and varied by exposure definition and study context. Higher magnitude of association between RAI and HV incidence were usually estimated when using more precise exposure definitions such time-varying RAI exposure, but the fraction of acts that are RAI were not associated with HIV incidence during VOICE. The results also showed differences between studies as the estimated association between reporting RAI at baseline and HIV incidence was much higher during RV 217 than during VOICE (aHR= 3.3 vs 1.1).

The prevalence of RAI among women at baseline was consistently higher than 15% across the three studies, but it decreased by more than 70% in VOICE and RV 217, and 25% in HVTN 907 at last follow-up, partly due to regression to the mean (30) or counselling of study participants, but not due to larger loss to follow-up among women reporting RAI. Interestingly, this decrease contrasts with the use of condoms with RVI and RAI during VOICE which remained steady over time despite counselling (Figure S9). Our analysis also showed that although few individuals continued practicing RAI throughout the studies, a non-negligible number (around 8%) reported initiating it during the first follow-up visit (Figure 2b), despite counselling. When available (VOICE, HVTN 907), RAI fraction appeared to remain stable over time and higher in VOICE (at risk women) than in HVTN 907 (FSW), which suggests that the risk of acquiring HIV during RAI among women still practicing RAI was consistently high over time.

In VOICE, the only study for which we could conduct all adjusted analysis, the estimates of the magnitude of association, albeit not necessarily statistically significant, were larger for exposure definitions that aimed measure precise and higher intensity RAI exposures (aHR=1.2 (0.9-1.6) using time varying covariate Cox proportional hazards model, and aHR=2.0 (1.3-3.1) among those always reporting RAI). The opposite was found for RV 217 where reporting RAI at baseline was associated with higher statistical associations (aHR=3.3; 1.6-6.8) than when defining exposures from follow-up data.

Our estimates are in line with pooled African estimates from Stannah at al. review, where the crude and adjusted measures of association were 1.2 (0.9−1.5) and 2.3 (0.8−6.4), respectively, based on 13 studies (17). In the review, finer definitions of RAI exposure were also not associated with higher increase in HIV incidence. The lower magnitude of the association during VOICE compared to RV 217 and HVTN could be partly explained by the higher incidence of HIV during the study (when risky behaviours are not needed to lead to HIV infection because of higher prevalence among partners, their association with HIV incidence is diminished), as well as less precise HIV testing algorithm compared to RV 217.

Limitations to our analysis are primarily due to the low HIV incidence during the RV 217 and HVTN 907 observational incidence studies. Only 12 incident infections occurred during the HVTN 907 study, and if 34 infections occurred in the Ugandan and Tanzanian RV 217 study sites, only 22 of them occurred among individuals reporting data during follow-up. When combined with the lower prevalence of RAI reported during follow-up, these sample sizes did not allow analysing the relationship between RAI practice and HIV incidence. The reliability of the estimated dates of HIV infection depended on the frequency and type of HIV tests used, which were rapid tests used monthly and every 6 months in the case of VOICE and HVTN, respectively, whereas RV 217 used RNA tests twice a week, minimising the risk of misclassification bias.

Despite efforts and improvements in the methods of data collection, RAI may still be not accurately reported by participants (18). This might be especially true here since the meaning of RAI is particularly ambiguous in several southern Africa local languages (18, 31). The VOICE and RV 217 studies relied on ACASI techniques that may yield higher RAI prevalence estimates, for example due to reduced social desirability biases (3), but this technique does not allow the interviewer checking how well the respondent understood the question. The second version of the VOICE behavioural questionnaire progressively administered within the South African and Ugandan sites during the trial did improve the translation of RAI practice questions in local languages (24) and reduced the possibility of misclassification biases. As a result, RAI prevalence at baseline was 8% relatively lower among those using this second questionnaire version, and this difference remained constant during the first year of follow-up (Figure S10). Therefore, the observed decrease in the cross-sectional RAI prevalence among VOICE participants may be partly due to the progressive improvement of translations of questions into local languages. However, the estimated associations between the different RAI exposures definitions and HIV incidence did not change when the HIV incidence analysis was conducted only among those using the second version of the VOICE behavioural ACASI questionnaire from baseline (23% of participants) (Figure S11). The reported fraction of acts that were RAI was not associated with HIV incidence during VOICE, which could be partly because the number of RVIs in the women was reported over only one week vs 3 months for RAI, leading to significant number of women not reporting RVI and only RAI, and a possible overestimation of the RAI fraction. Another possible source of misclassification bias is due to the RAI prevalence exposure in the RV 217 analysis being assumed to cover the 6-months period between two behavioural assessments, whereas the question about RAI practice only covered the past 3 months before each assessment.

The strength of our analysis is that it benefitted from the context of the HIV trials (VOICE) or observational incidence studies (RV 217, HVTN 907), where different RAI exposure definitions and their association with HIV incidence was evaluated using frequent behavioural surveys and HIV tests, increasing the robustness and accuracy of results. As we have seen during the analysis, the practice of RAI is not stable over time, with many women reporting initiating and stopping RAI over time.

When based on cross-sectional data, only HIV prevalence by RAI status at the time of data collection can be studied, leading to potential reverse causality issues (past HIV infection could explain changes in behaviours). Our analyses of the HIV incidence were adjusted on several important cofactors such as number of partners or condom use, and other higher risk practices such as sex work and injecting drugs, leading to more reliable estimates of association. Condom use was available at last sex (or RVI), but also at last RAI specifically for VOICE and RV 217. In VOICE, the levels of condom use at last RVI and last RAI were similar, whilst during RV 217 condom use was less frequent among RAI+ women compared to RVI-only women (which was accounted for by our statistical models), but condom use by RAI+ women during RAI was similar to during RVI (Table S1). However, our models could not be controlled for the HIV status of the participants’ male partners which were not known. It is possible that the lack of association between RAI and HIV incidence could be confounded by different distributions of demographic factors and HIV status (including viral load suppression) of the clients being able to afford a paid RAI (in Africa the price of RAI with a sex worker is much more expensive than a RVI (13)).

## Conclusions

Estimating the association between RAI and HIV acquisition is challenging, due to misclassification biases and uncertainties around the time of HIV infection, but our analysis shows that RAI increases the risk of HIV even among women already at high risk of HIV acquisition, and that this practice must be more systematically measured and better understood. Future serodiscordant couple studies collecting behavioural data using anonymous, regular, and precise RAI exposure measurements, as well as performing regular HIV tests and biomarkers may help obtaining more accurate pictures of the actual RAI practice of their participants. This will allow better assessment of the association between RAI practice and HIV risk which would subsequently help evaluating the impact of HIV prevention tools targeting both vaginal and anal sites.

## Data Availability

The data and material analysed can be obtained following specific procedures.

## Acknowledgments

The authors would like to express heartfelt appreciation to Dr Peter Anton from the University of California, Los Angeles and Dr Edith Swann from the US national institute of health for their support and comments on the project. We would also like to thank Dr Mike Chirenje from the University of Zimbabwe and Dr Barbra Richardson from the University of Washington for providing insight on the VOICE data.

## References

1. McBride KR, Fortenberry JD. Heterosexual anal sexuality and anal sex behaviors: a review. J Sex Res. 2010;47(2):123–36.

2. Voeller B. AIDS and heterosexual anal intercourse. Arch Sex Behav. 1991;20(3):233–76.

3. Owen BN, Elmes J, Silhol R, Dang Q, McGowan I, Shacklett B, et al. How common and frequent is heterosexual anal intercourse among South Africans? A systematic review and meta-analysis. J Int Aids Soc. 2017;19(1):1–14.

4. Baggaley RF, White RG, Boily MC. HIV transmission risk through anal intercourse: systematic review, meta-analysis and implications for HIV prevention. Int J Epidemiol. 2010;39(4):1048–63.

5. Halperin DT. Heterosexual anal intercourse: prevalence, cultural factors, and HIV infection and other health risks, Part I. AIDS patient care and STDs. 1999;13(12):717–30.

6. Owen BN, Brock PM, Butler AR, Pickles M, Brisson M, Baggaley RF, et al. Prevalence and Frequency of Heterosexual Anal Intercourse Among Young People: A Systematic Review and Meta-analysis. Aids Behav. 2015;19(7):1338–60.

7. Baggaley RF, Dimitrov D, Owen BN, Pickles M, Butler AR, Masse B, et al. Heterosexual anal intercourse: a neglected risk factor for HIV? Am J Reprod Immunol. 2013;69 Suppl 1:95–105.

8. Morhason-Bello IO, Kabakama S, Baisley K, Francis SC, Watson-Jones D. Reported oral and anal sex among adolescents and adults reporting heterosexual sex in sub-Saharan Africa: a systematic review. Reprod Health. 2019;16(1):48.

9. Reynolds GL, Fisher DG, Rogala B. Why women engage in anal intercourse: results from a qualitative study. Arch Sex Behav. 2015;44(4):983–95.

10. Owen BN, Baggaley RF, Maheu-Giroux M, Elmes J, Adimora AA, Ramirez C, et al. Patterns and Trajectories of Anal Intercourse Practice Over the Life Course Among US Women at Risk of HIV. J Sex Med. 2020;17(9):1629–42.

11. Boily MC, Baggaley RF, Wang L, Masse B, White RG, Hayes RJ, et al. Heterosexual risk of HIV-1 infection per sexual act: systematic review and meta-analysis of observational studies. Lancet Infect Dis. 2009;9(2):118–29.

12. Baggaley RF, Owen BN, Silhol R, Elmes J, Anton P, McGowan I, et al. Does per-act HIV-1 transmission risk through anal sex vary by gender? An updated systematic review and meta-analysis. American Journal of Reproductive Immunology. 2018;80(5).

13. Maheu-Giroux M, Baral S, Vesga JF, Diouf D, Diabaté S, Alary M, et al. Anal Intercourse Among Female Sex Workers in Côte d’Ivoire: Prevalence, Determinants, and Model-Based Estimates of the Population-Level Impact on HIV Transmission. Am J Epidemiol. 2017;187(2):287–97.

14. McGowan I, Taylor DJ. Heterosexual Anal Intercourse Has the Potential to Cause a Significant Loss of Power in Vaginal Microbicide Effectiveness Studies. Sex Transm Dis. 2010;37(6):361–4.

15. Masse BR, Boily MC, Dimitrov D, Desai K. Efficacy dilution in randomized placebo-controlled vaginal microbicide trials. Emerging themes in epidemiology. 2009;6:5.

16. Boily MC, Dimitrov D, Karim SSA, Masse B. The future role of rectal and vaginal microbicides to prevent HIV infection in heterosexual populations: implications for product development and prevention. Sex Transm Infect. 2011;87(7):646–53.

17. Stannah J, Silhol R, Elmes J, Owen B, Shacklett BL, Anton P, et al. Increases in HIV Incidence Following Receptive Anal Intercourse Among Women: A Systematic Review and Meta-analysis. Aids Behav. 2019.

18. Duby Z, Hartmann M, Mahaka I, Munaiwa O, Nabukeera J, Vilakazi N, et al. Lost in Translation: Language, Terminology, and Understanding of Penile-Anal Intercourse in an HIV Prevention Trial in South Africa, Uganda, and Zimbabwe. J Sex Res. 2015:1–11.

19. Peebles K, van der Straten A, Palanee-Phillips T, Reddy K, Hillier SL, Hendrix CW, et al. Brief Report: Anal Intercourse, HIV-1 Risk, and Efficacy in a Trial of a Dapivirine Vaginal Ring for HIV-1 Prevention. Journal of acquired immune deficiency syndromes (1999). 2020;83(3):197–201.

20. Owen BN, Baggaley RF, Maheu-Giroux M, Elmes J, Adimora AA, Ramirez C, et al. Longitudinal determinants of anal intercourse among women with, and without HIV in the United States. Bmc Womens Health. 2022;22(1).

21. Robb ML, Eller LA, Kibuuka H, Rono K, Maganga L, Nitayaphan S, et al. Prospective Study of Acute HIV-1 Infection in Adults in East Africa and Thailand. N Engl J Med. 2016;374(22):2120–30.

22. Palanee-Phillips T, Schwartz K, Brown ER, Govender V, Mgodi N, Kiweewa FM, et al. Characteristics of Women Enrolled into a Randomized Clinical Trial of Dapivirine Vaginal Ring for HIV-1 Prevention. Plos One. 2015;10(6).

23. Marrazzo JM, Ramjee G, Richardson BA, Gomez K, Mgodi N, Nair G, et al. Tenofovir-Based Preexposure Prophylaxis for HIV Infection Among African Women. Obstet Gynecol Surv. 2015;70(7):444–6.

24. Marrazzo JM, Ramjee G, Richardson BA, Gomez K, Mgodi N, Nair G, et al. Tenofovir-Based Preexposure Prophylaxis for HIV Infection among African Women. New Engl J Med. 2015;372(6):509–18.

25. Robb ML, Eller LA, Kibuuka H, Rono K, Maganga L, Nitayaphan S, et al. Prospective Study of Acute HIV-1 Infection in Adults in East Africa and Thailand. New Engl J Med. 2016;374(22):2120–30.

26. Deschamps MM, Metch B, Morgan CA, Zorilla CD, Donastorg Y, Swann E, et al. Feasibility of Identifying a Female Sex Worker Cohort at High Risk of HIV Infection in the Caribbean for HIV Vaccine Efficacy Trials: Longitudinal Results of HVTN 907. Jaids-J Acq Imm Def. 2016;71(1):70–7.

27. Cox DR. Regression Models and Life-Tables. J R Stat Soc B. 1972;34(2):187-+.

28. Fisher LD, Lin DY. Time-dependent covariates in the Cox proportional-hazards regression model. Annu Rev Public Health. 1999;20:145–57.

29. R Core Team. R: A Language and Environment for Statistical Computing Vienna, Austria 2013 [Available from: http://www.R-project.org.

30. Barnett AG, van der Pols JC, Dobson AJ. Regression to the mean: what it is and how to deal with it. Int J Epidemiol. 2004;34(1):215–20.

31. Gorbach PM, Mensch BS, Husnik M, Coly A, Masse B, Makanani B, et al. Effect of Computer-Assisted Interviewing on Self-Reported Sexual Behavior Data in a Microbicide Clinical Trial. Aids Behav. 2013;17(2):790–800.

