## SupplementFigures for "The association between heterosexual anal intercourse and HIV acquisition in three prospective cohorts of women"

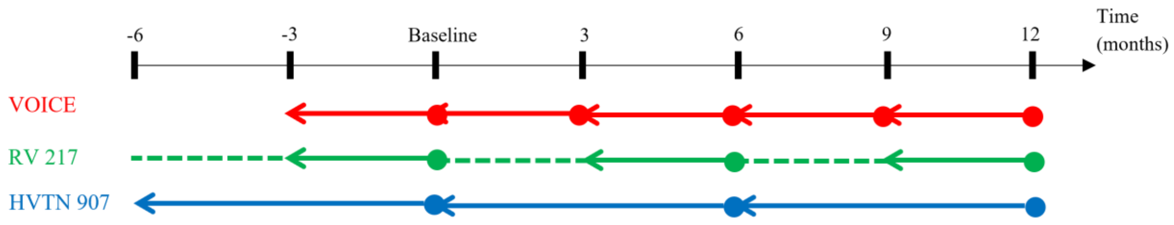


**Figure S1.** Reporting of RAI prevalence data during VOICE (red), RV 217 (green), and HVTN 907 (blue) longitudinal surveys. Each dot represents the time of the baseline or follow-up behavioural survey, whereas plain arrows represent the recall periods over which RAI prevalence is reported. The green dashed line represents periods of RV 217 survey which were not covered by the behavioural questionnaire, and our analysis assumes that the participants' report of RAI practice over their past six months was the same as over the past 3 months.

Table S1: Summary of characteristics of the study participants at baseline

| | VOICE | RV 217 | HVTN-907 |
|---|---|---|---|
| Country | South Africa: 81%<br>Uganda: 6%<br>Zimbabwe: 13% | Kenya: 60%<br>Uganda: 40% | Dominican Republic: 32%<br>Haiti: 44%<br>Puerto Rico: 24% |
| Trial arm | Intervention: 60%<br>Placebo: 40% | NA | NA |
| Age | <25 yrs: 51%<br>≥25 yrs: 49% | <25 yrs: 62%<br>≥25 yrs: 38% | <25 yrs: 46%<br>≥25 yrs: 54% |
| Sex work | Yes: 6%[a]<br>No: 94% | Yes: 82%[b]<br>No: 18% | Yes: 100%[c]<br>No: 0% |
| Injection drug use | NA | Ever: 8%, including current: 2% | In the past 6 months: 3%<br>Not in the past 6 months: 97% |
| Number of partners | 0: 2%[d]<br>1: 77%<br>2: 16%<br>3+: 5% | 0: 7% [e]<br>1-2: 37%<br>3-10: 32%<br>10+: 24% | 0-10: 15% [f]<br>11-100: 22%<br>101-1000: 51%<br>1000+: 12% |
| Number of sex acts | 0: 28%[g]<br>1: 16%<br>2: 20%<br>3: 16%<br>4+: 20% | 0-2: 6% [h]<br>3-10: 15%<br>10-100: 70%<br>101+: 9% | NA |
| Condom use at last RVI | Among RVI-only women: 71%<br>Among RAI+ women: 73% | At last intercourse (not reported if RAI or RVI)<br>a) with a steady partner:<br>RVI-only women: 45%<br>RAI+ women: 43%<br>b) with a casual partner:<br>RVI-only: 68%<br>RAI+: 48%<br>c) with a client:<br>RVI-only: 72%<br>RAI+: 53%<br>d) with another[i] type of partner:<br>RVI-only: 63%<br>RAI+: 49% | Among RVI-only women: 79%<br>Among RAI+ women: 73% |
| Condom use at last RAI | 69% | With a steady partner: 39%<br>With a casual partner: 43%<br>With a client: 47%<br>With another[i] type of partner: 47% | NA |

[a] Defined as having received money or good in exchange for sex in the past year
[b] Defined as having had a commercial partner in the past 3 months
[c] Defined as having received money or good in exchange for sex in the past 6 months
[d] Defined in the past 3 months
[e] Defined as the number of partners they had condomless sex with in the past 3 months
[f] Defined as the number of clients seen in the past 6 months
[g] Defined in the past week
[h] Defined in the past 3 months
[i] "other" partners include boss/work supervisor, teacher, other authority figure and relative other than spouse, student, employee/work subordinate, or rapist.

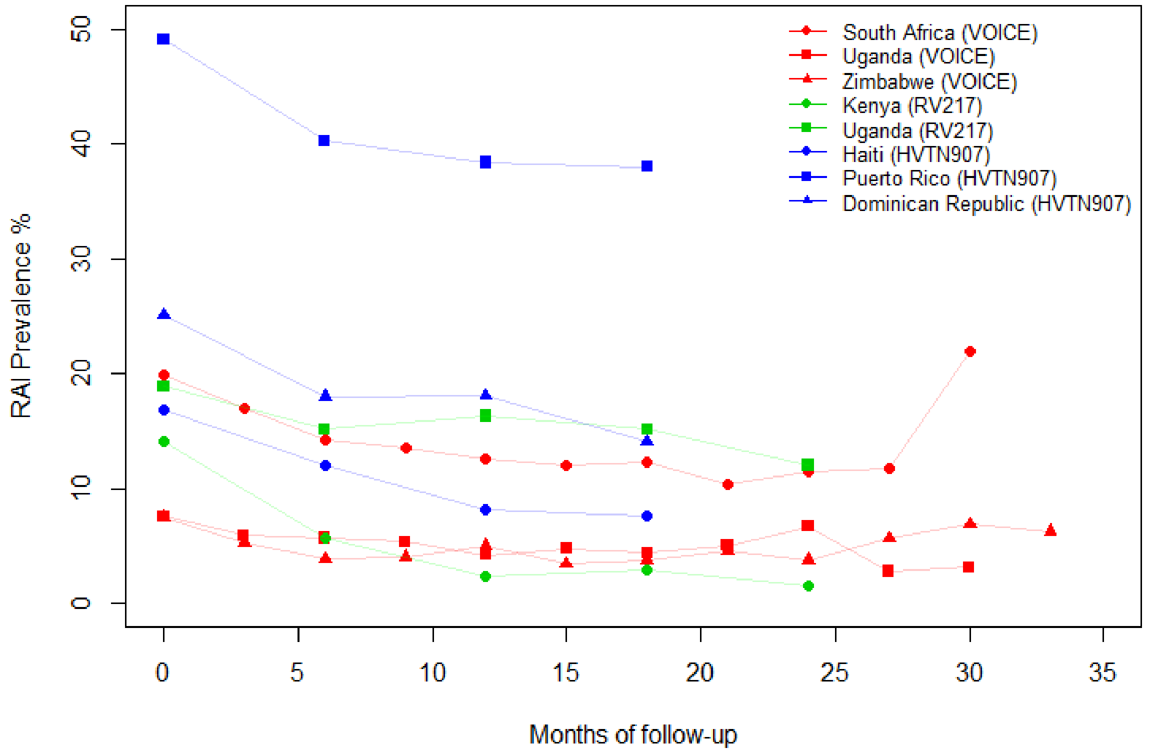


**Figure S2.** Cross-sectional RAI prevalence for the three longitudinal studies under consideration (VOICE, RV 217, and HVTN 907), by participants country of residence. Differences between RAI prevalence in Uganda as reported by the women from VOICE and RV 217 (red and green lines) are partly due to the fact that the RV 217 study enrolled high-risker risk women.

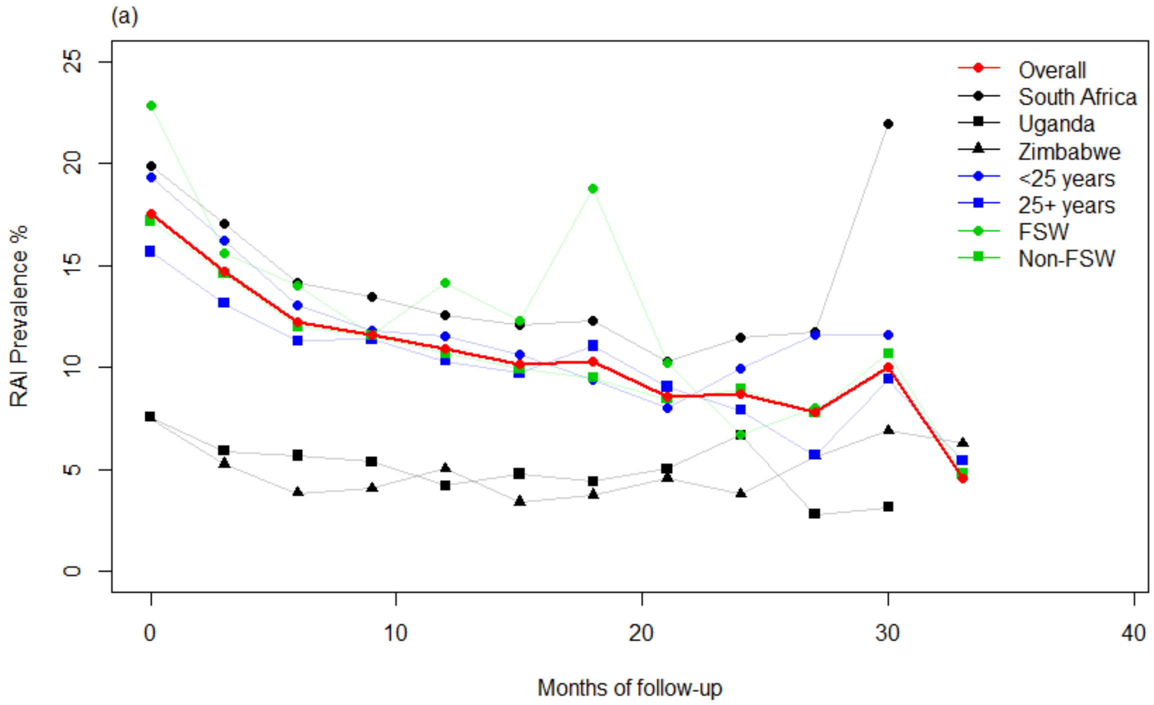
(a)
RAI Prevalence %
Months of follow-up
0
5
10
15
20
25
0
10
20
30
40
Overall
South Africa
Uganda
Zimbabwe
<25 years
25+ years
FSW
Non-FSW

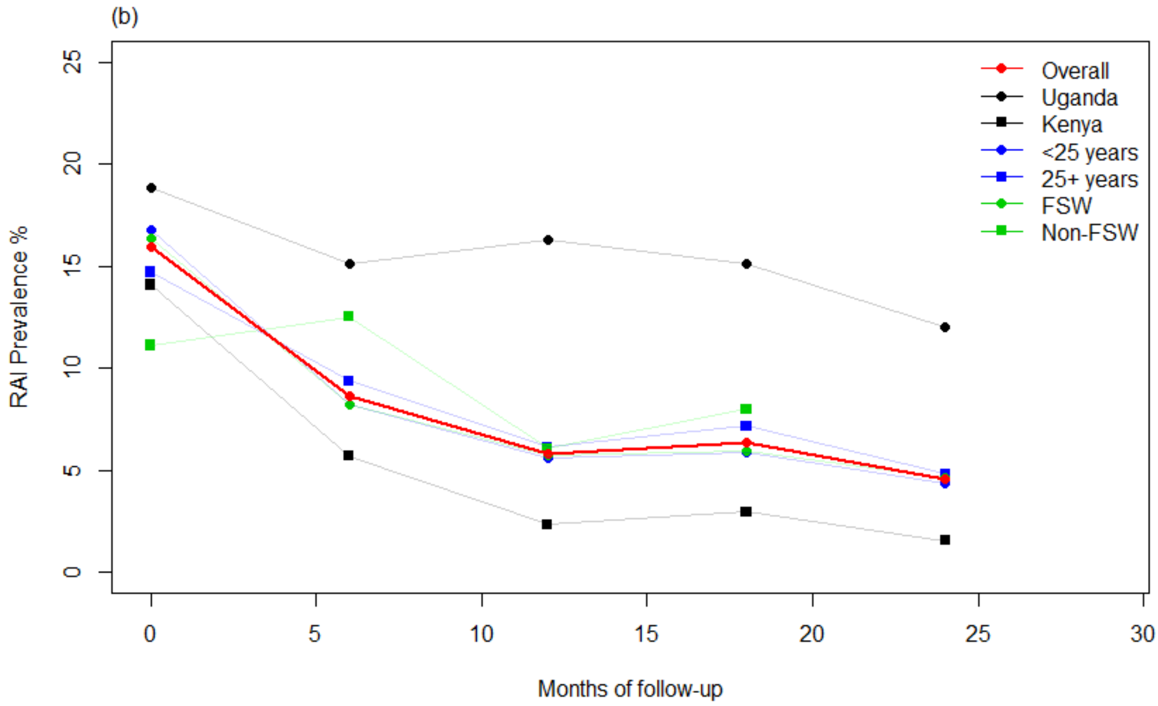
(b)
RAI Prevalence %
Months of follow-up
0
5
10
15
20
25
0
5
10
15
20
25
30
Overall
Uganda
Kenya
<25 years
25+ years
FSW
Non-FSW

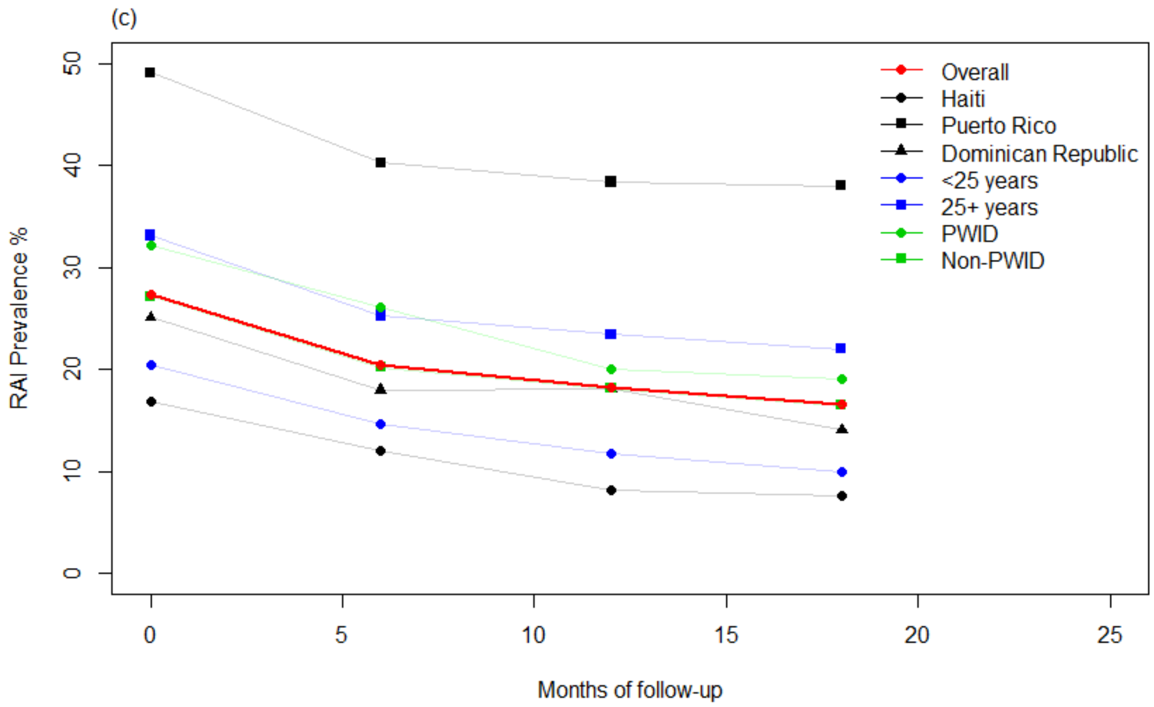


**Figure S3:** Cross-sectional RAI prevalence during a) MTN-003 VOICE, b) RV 217, c) HVTN 907 studies, stratified by individual characteristics.

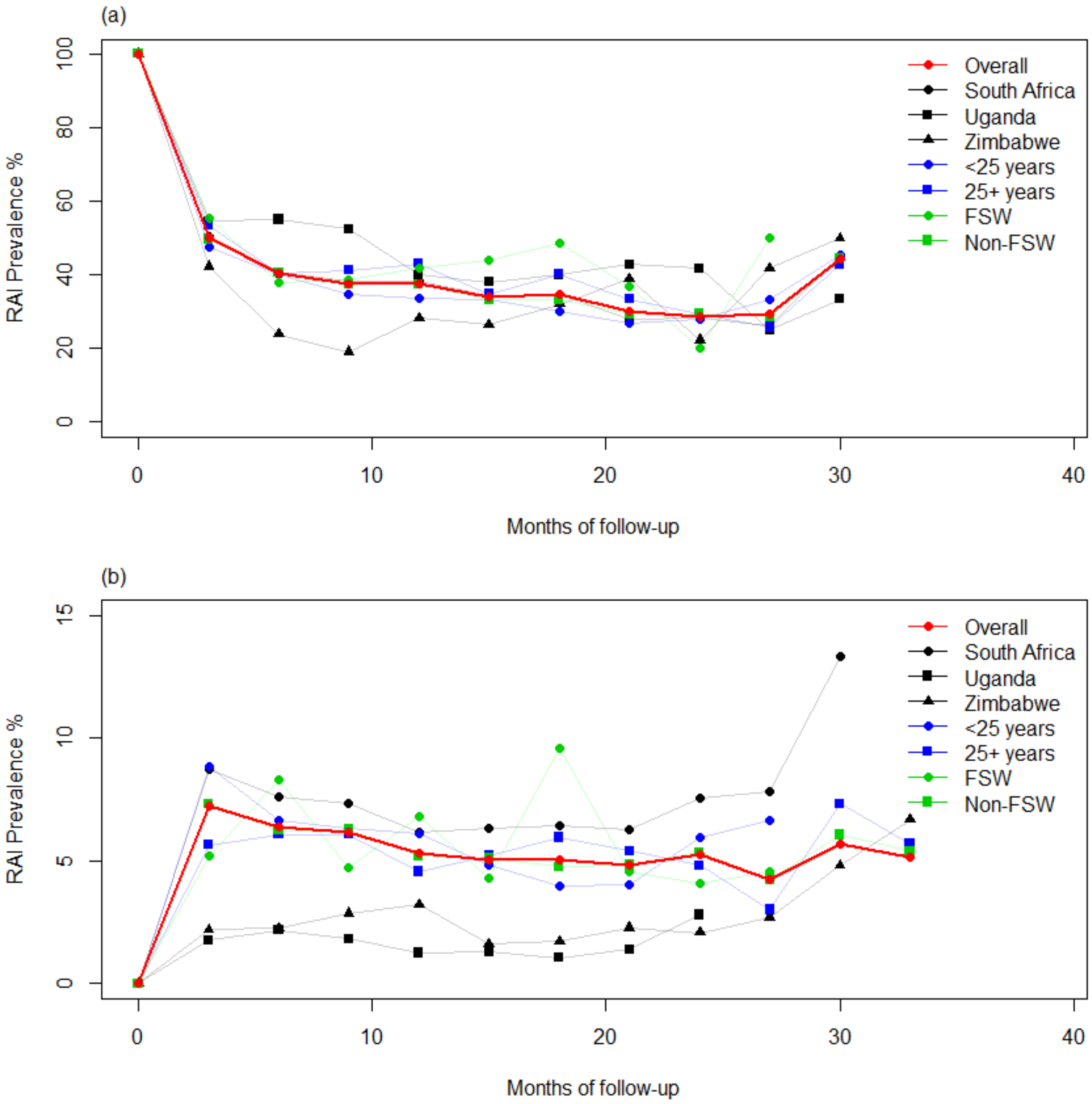


**Figure S4:** Cross-sectional RAI prevalence during VOICE among (a) RAI+ women and (b) RVI-only women at baseline, stratified by individual characteristics.

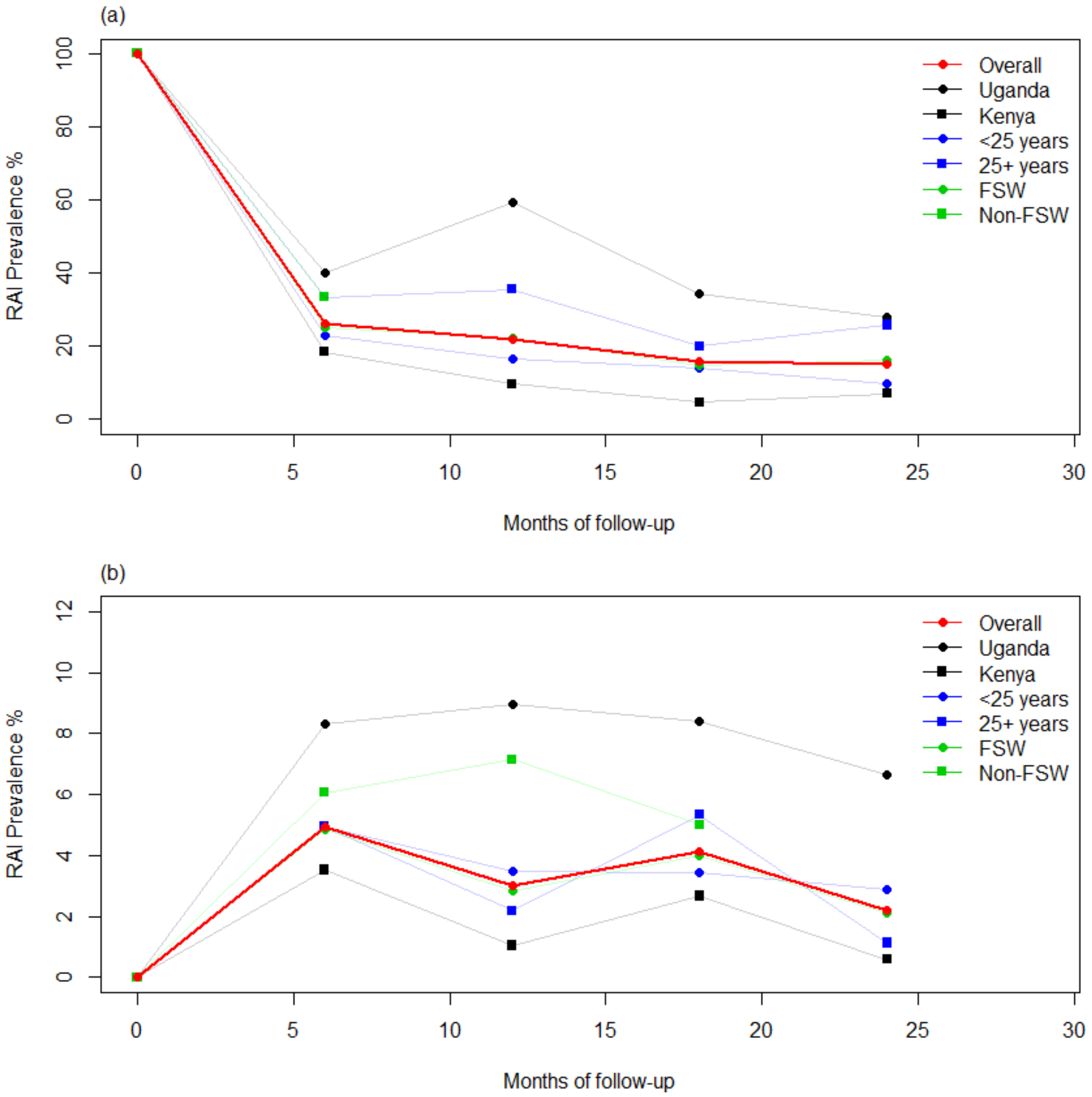


**Figure S5:** Cross-sectional RAI prevalence during RV 217 among (a) RAI+ women and (b) RVI-only women at baseline, stratified by individual characteristics.

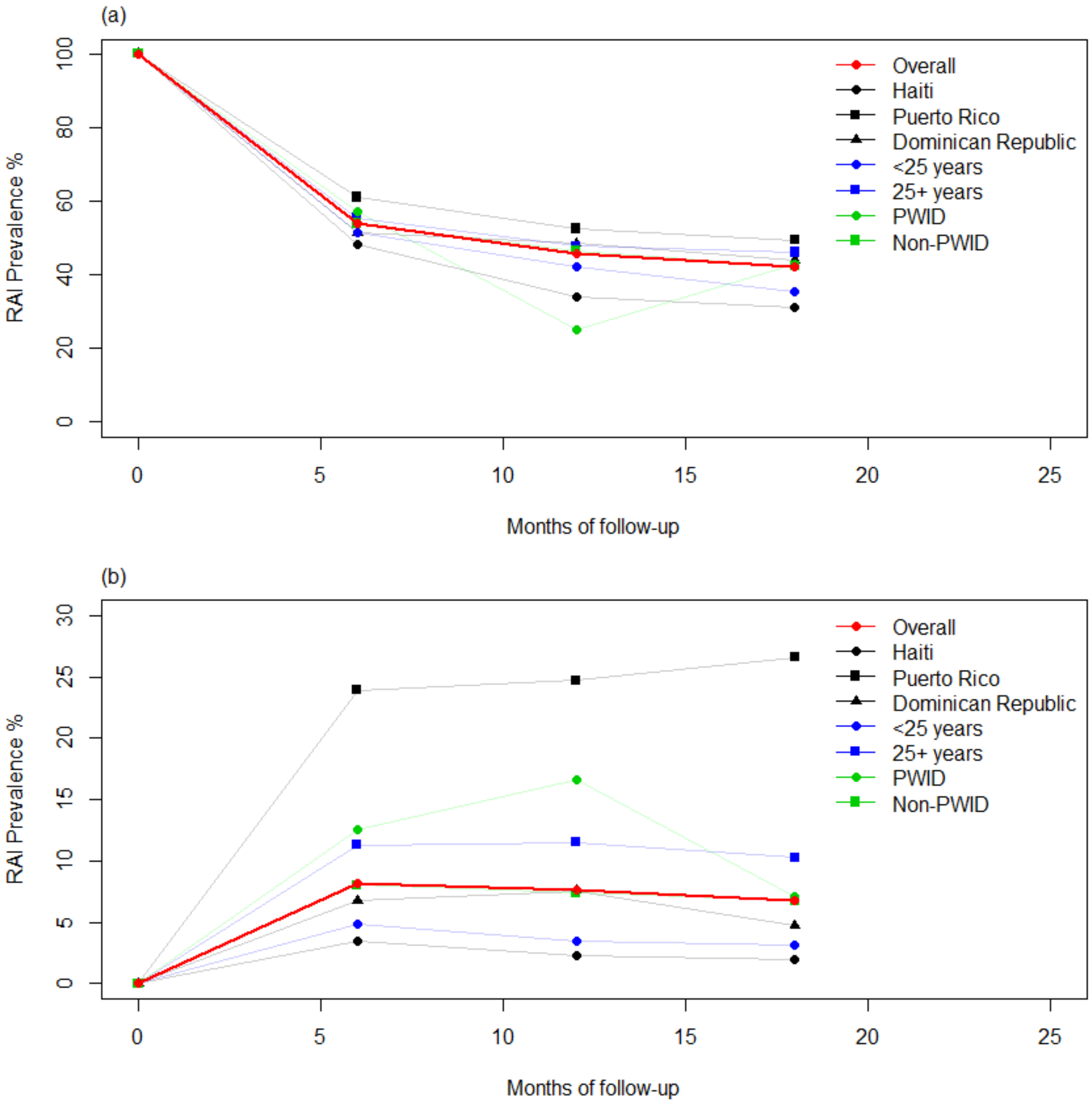


**Figure S6:** Cross-sectional RAI prevalence during HVTN 907 among (a) RAI+ women and (b) RVI-only women at baseline, stratified by individual characteristics.

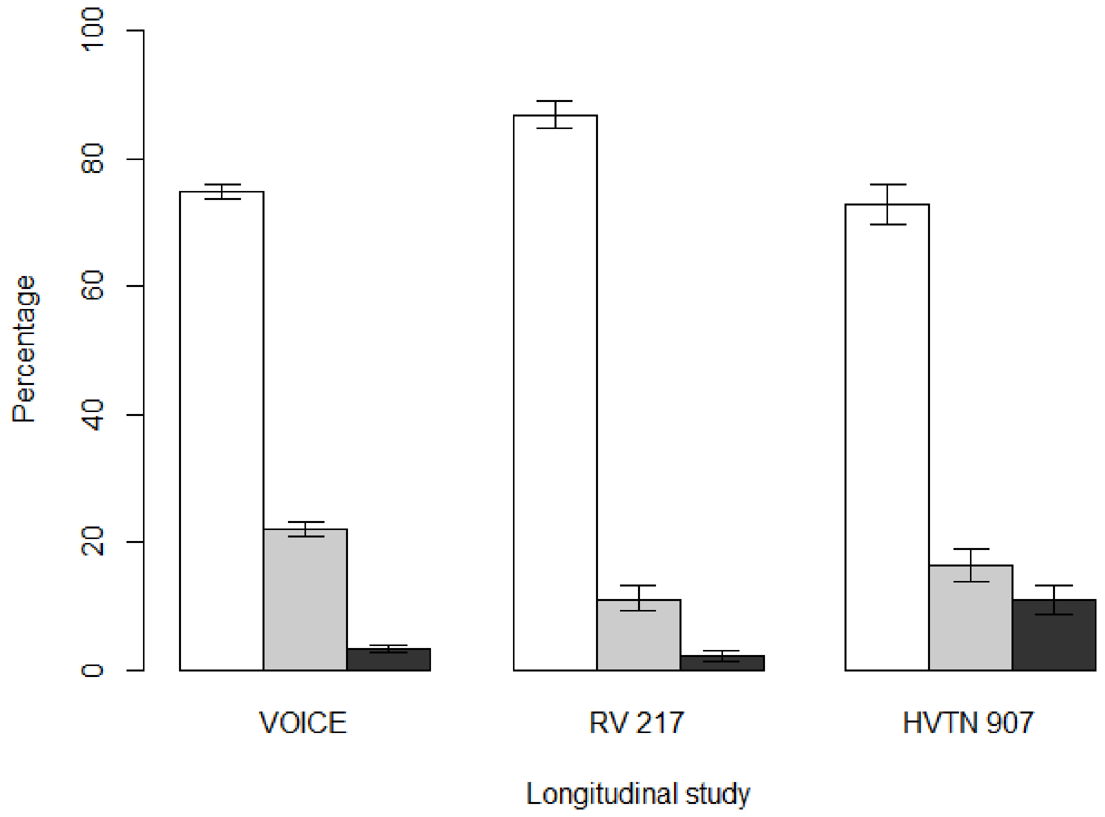


**Figure S7.** Stability of RAI practice: Proportion of individuals of each study that never (white bars), sometimes (grey bars), or always (dark bars) reported RAI during follow-up. Errors bars represent 95%CI of data.

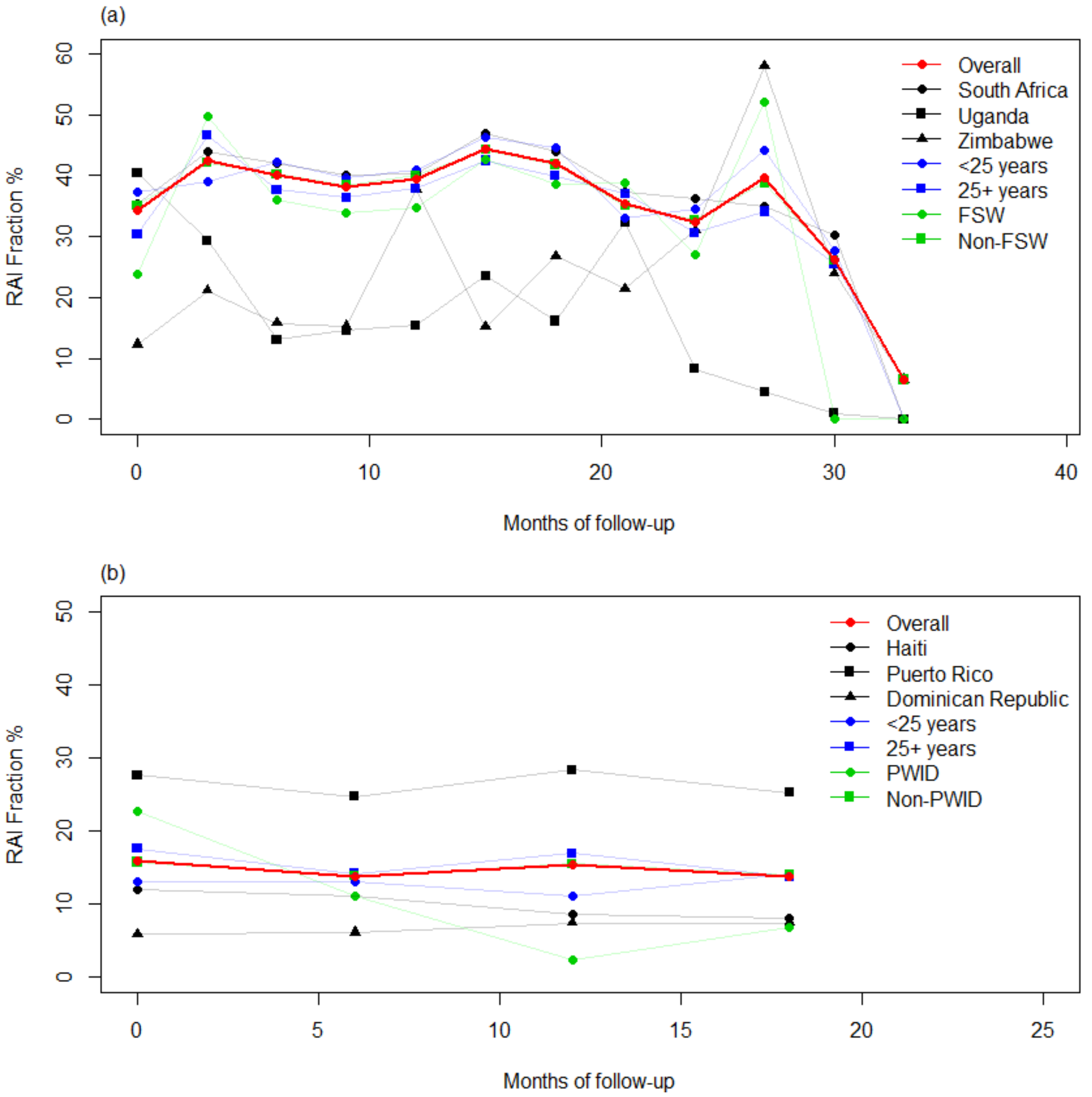


**Figure S8:** Cross-sectional RAI fraction (% sex acts that are anal among individuals reporting RAI) during VOICE and HVTN 907, calculated among subpopulations.

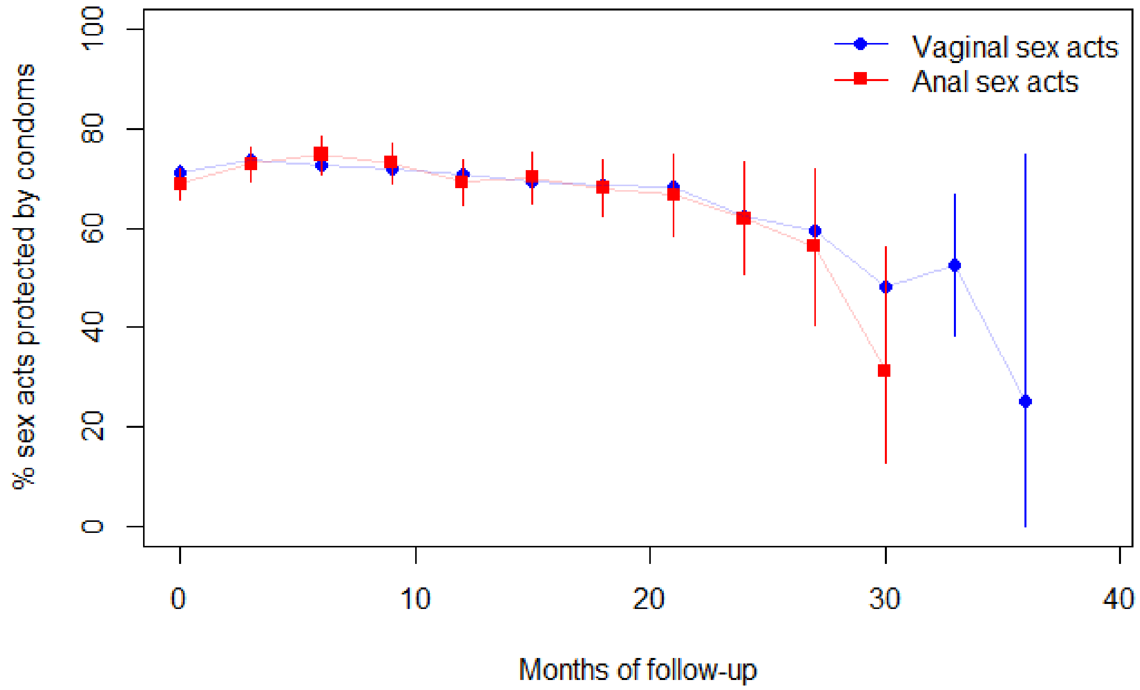


**Figure S9:** Cross-sectional proportion of last vaginal (blue) and anal (red) sex acts during which condoms were used, as reported by VOICE participants. Errors bars represent 95%CI of data.

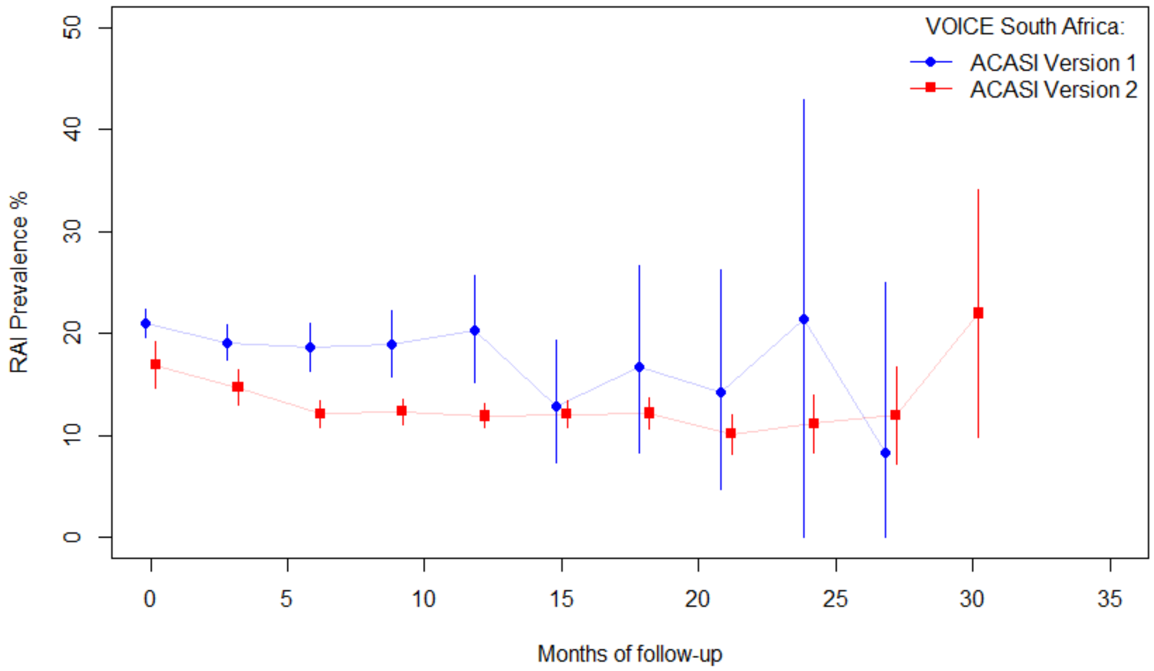


**Figure S10:** Cross-sectional RAI prevalence during VOICE by ACASI questionnaire version, reported by South African participants. Errors bars represent 95%CI of data.

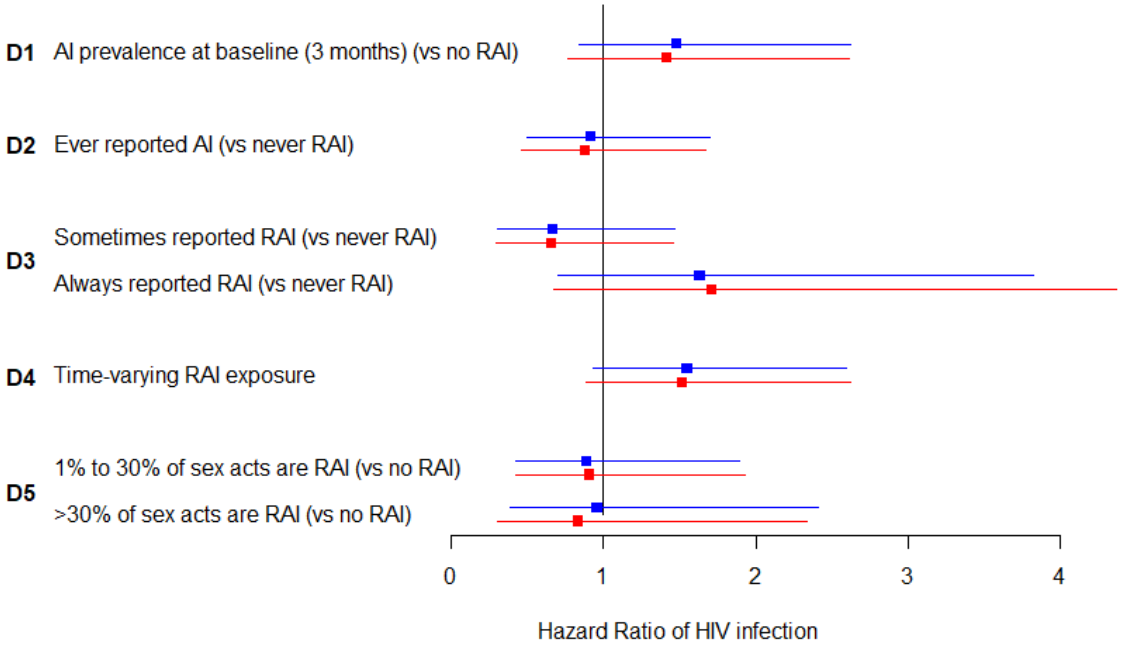


**Figure S11:** Increase in HIV incidence among women practicing RAI during VOICE, Unadjusted (blue) and adjusted (red) estimates of the association between different exposures of RAI and HIV incidence during VOICE, only among those who used the second version of the ACASI questionnaire (23% of participants). Confounding variables include country, age, trial arm, sex work in the prior year, number of partners, and condom use at last vaginal sex. Errors bars represent 95%CI of estimates.
